# Predictive performance of SCORE2 among people with depression and anxiety disorder

**DOI:** 10.1101/2025.04.08.25325451

**Authors:** Shinya Nakada, Carlos Celis-Morales, Jill P Pell, Frederick K Ho

## Abstract

**Background:** It is not known how well SCORE2, a cardiovascular disease (CVD) risk score commonly used in Europe, performs among people with depression or anxiety disorder.

**Methods:** Among UK Biobank participants, prevalent depression and anxiety disorder were ascertained through self-report and hospital admission and primary care data. Prediction of 10-year CVD risk using SCORE2 was evaluated among those with and without depression/anxiety disorder by discrimination and calibration measures. Recalibration impact was assessed in terms of the distributions of individuals across predicted risk categories; and potentially preventable cases, estimated by dividing the additional number of individuals in the intermediate or higher predicted risk category by the number needed to treat (NNT) using statins.

**Results:** Among 34,228 participants with depression/anxiety disorder, 3,076 (9.0%) had CVD events over a 10-year follow-up period. SCORE2 had a lower C-index (0.695, 95% CI 0.684–0.706) among those with depression/anxiety compared to those without (0.710, 95% CI 0.706–0.713). The calibration plot indicated that predicted risk was lower than observed (E/O ratio 0.782, 95% CI 0.746–0.818; calibration slope 0.899, 95% CI 0.868–0.930) among those with depression/anxiety disorder. After recalibration, 5,529 (38%) of those aged ≥50 years and 1,464 (23%) of those aged <50 years with these mental health conditions were reclassified upward to the intermediate or higher risk category. Dividing these by NNT using statins yielded 191 (11.7%) and 51 (27.4%) additional preventable CVD events among those aged ≥50 and <50 years, respectively.

**Conclusions:** SCORE2 could underestimate 10-year CVD risk among people with depression or anxiety disorder, potentially leading to CVD events that could have been prevented with statin therapy.

## Background

One in every five women and one in every ten men experience depression or anxiety disorder during their lifetime [1]. People with these common mental health conditions are at a greater risk of cardiovascular disease (CVD) [2], which contributes to their shorter life expectancy [3]. Assessing the risk of CVD is of particular importance in this high-risk population.

The Systematic COronary Risk Evaluation 2 (SCORE2) is a risk assessment tool of incident CVD over 10 years developed for European countries [4]. SCORE, the predecessor of SCORE2, was the most widely used assessment tool in European countries [5]. Predicted risk guides primary prevention through the management of CVD risk factors, such as low-density lipoprotein cholesterol (LDL-c), in the general populations, in line with the European Society of Cardiology guidelines [6].

However, it is unknown how accurately SCORE2 predicts CVD risk in people living with common mental health conditions including depression and anxiety disorder. Given their high CVD rate, distinct behavioural, physical and metabolic characteristics and use of psychiatric drugs [7–10], the SCORE2 predictive performance could differ from that in general populations and, if so, this may be contributing to the excess burden of CVD on this population, particularly in European countries.

Therefore, our study aimed to validate SCORE2 among middle- and old-aged people with depression or anxiety disorder and assess the potential impact of recalibration.

## Methods

### Study design and participants

This was an external validation study of SCORE2 using the UK Biobank cohort data. UK Biobank recruited over 500,000 participants between 2007 and 2010 from the general population, aged 40 to 69 years [11]. Participants were asked to undergo a baseline assessment at one of 22 centres across England, Scotland, and Wales and have been followed up via linkage to routine National Health Service databases. We excluded participants who had a history of CVD or diabetes or had missing values for any of the predictor variables.

We identified participants who had a depression or anxiety disorder diagnosis prior to the baseline ascertained through the self-completed baseline questionnaire or identified via retrospective linkage to hospital admission and primary care data. We defined depression and anxiety disorder as the International Classification of Diseases, 10th revision (ICD-10) codes of F32–33 and F40–43, respectively. Participants who did not have either of these conditions were also included as the comparison group.

### Outcome

The outcome was incident CVD in line with the SCORE2 outcome. Non-CVD deaths were treated as competing events. The ICD-10 codes used to ascertain CVD events are provided in the supplementary Table 1. The death certificate data were available up to November 2022 and were obtained from the National Health Service Information Centre and for England and Wales and from the National Health Service Central Register Scotland for Scotland. The hospital admission data were available up to October 2022 for England and May 2022 for Wales from the Health Episode Statistics and up to August 2022 for Scotland from the Scottish Morbidity Records. Our study restricted the follow-up time to 10 years. Therefore, participants were censored at the end of the 10-year follow-up period or at the date of incident CVD or non-CVD death if either occurred earlier.

### Predictors

The SCORE2 predictors were: age, current smoking, systolic blood pressure (SBP), total cholesterol (TC), and high-density lipoprotein cholesterol (HDL). Age and current smoking status were self-reported by participants using a touchscreen questionnaire at the baseline assessment and treated as a continuous variable and a binary variable (yes/no), respectively. SBP was measured by a trained nurse at the baseline assessment and treated as a continuous variable. TC and HDL-c were measured on baseline blood samples and treated as continuous variables. Details of these assay performances are available in the protocol [12].

### Statistical analyses

There were three stages to our analyses: evaluating the SCORE2 performance, recalibrating the model, and assessing the impact of recalibration. The predictive performance of 10-year CVD risk was evaluated for those with depression or anxiety disorder and those without these conditions by applying the baseline survival and coefficients obtained from the published SCORE2 study [4]. We specified low-risk regions to calibrate risk estimates.

Discrimination was assessed by Harrell’s C-indices; calibration was assessed by calibration slopes, the expected/observed events (E/O) ratio, and calibration plots. 95% confidence intervals (CIs) were derived from the variance of these statistics. The calibration slopes were derived by fitting a Cox proportional hazard model with a linear predictor. The observed event probability of the E/O ratio was estimated using the Kaplan-Meier estimator. The model recalibration was conducted for those with depression or anxiety disorder by updating the baseline survival at 10 years in our data and checked by calibration plots.

Lastly, the age-specific impact of the recalibrated models was assessed in two stages. Firstly, the distribution of individuals across different predicted risk categories was compared between the original and recalibrated SCORE2. Predicted risk categories were defined as low, intermediate, and high, using cut-off values of <5%, 5% to <10%, ≥10% for those aged ≥50 years and <2.5%, 2.5% to <7.5%, ≥7.5% for those aged <50 years, in line with the European Society of Cardiology guideline. Secondly, the difference in the number of individuals assigned to the intermediate or higher risk category was divided by the number needed to treat (NNT) using statin therapy to estimate potentially preventable cases.

Because NNT is an outcome-specific measure, we calculated NNT based on 10-year risk of relevant CVD outcomes (CVD death, incident coronary heart disease, and incident stroke) of the West of Scotland Coronary Prevention Study [13]. This trial was specifically selected because of its focus on CVD primary prevention and the publicly available 10-year follow-up data.

Subgroup analyses were conducted for depression and anxiety disorder separately by repeating all procedures. All analyses were complete case analyses given the small proportions of missing values of all data (2.4%). However, we described the characteristics of those included and excluded due to missing values to confirm whether there were any differences. We confirmed a priori that both the numbers of events and non-events exceeded the conventional rule of 100 [14]. All analyses were conducted using R (version 3.5.3) package survival (version 3.2-7).

### Ethics Statement

All procedures contributing to this work comply with the ethical standards of the relevant national and institutional committees on human experimentation and with the Helsinki Declaration of 1975, as revised in 2013. UK Biobank was approved by the North-West Multi-Centre Research Ethics Committee (Ref: 11/NW/0382). This work was conducted under the UK Biobank application number 71392.

### Consent Statement

All participants provided written informed consent.

## Results

In total, 401,453 participants were included in the analyses after excluding those who had a history of CVD or diabetes or missing values in predictors (Supplementary Figure 1). Among 34,228 participants with existing depression or anxiety disorder, 3,076 (9.0%) had CVD events over a mean follow-up of 9.6 years; among 367,225 participants without these conditions, 27,355 (7.4%) had CVD events over a mean follow-up of 9.7 years (Table 1).

**Table 1.** Characteristics of participants with depression or anxiety disorder.

|  | Participants without depression or anxiety disorder |  |  | Participants with depression or anxiety disorder |  |  |
| --- | --- | --- | --- | --- | --- | --- |
|  | Total | Non-CVD | CVD | Total | Non-CVD | CVD |
|  | N = 367,225 | N = 339,870 | N = 27,355 | N = 34,228 | N = 31,152 | N = 3,076 |
| Age (years), mean (SD) | 56.5 (8.11) | 56.1 (8.09) | 61.4 (6.62) | 55.5 (7.87) | 55.1 (7.83) | 59.8 (6.98) |
| Smoking, N (%) | 36,515 (9.94) | 32,138 (9.46) | 4,377 (16.00) | 5,607 (16.38) | 4,876 (15.65) | 731 (23.76) |
| Systolic blood pressure (mmHg), mean (SD) | 138.1 (18.60) | 137.6 (18.42) | 145.2 (19.42) | 135.1 (18.31) | 134.5 (18.05) | 141.6 (19.66) |
| Total cholesterol (mmol/l), mean (SD) | 5.7 (1.12) | 5.7 (1.11) | 5.6 (1.23) | 5.8 (1.16) | 5.8 (1.14) | 5.7 (1.31) |
| HDL-c (mmol/l), mean (SD) | 1.5 (0.38) | 1.5 (0.38) | 1.4 (0.37) | 1.4 (0.38) | 1.5 (0.38) | 1.4 (0.38) |

CVD, cardiovascular disease; N, number; SD, standard deviation; HDL-c, high density lipoprotein cholesterol

Those with depression or anxiety disorder were more likely to be a current smoker (16.4% vs 9.9%) and had lower SBP (135.1 vs 138.1 mmHg) than those without conditions. Overall, those who developed CVD were older and more likely to be a current smoker and have higher SBP.

SCORE2 had a lower C-index (0.695, 95% CI: 0.684–0.706) among those with depression or anxiety disorder compared to those without (0.710, 95% CI: 0.706–0.713) (Table 2). The E/O ratio (0.782, 95% CI: 0.746–0.818) and calibration slope (0.899, 95% CI: 0.868–0.930) were also lower, consistent with the calibration plot showing the upward deviation of the curve from the diagonal (Figure 1). However, after recalibration, the calibration curve appeared to lie on the diagonal, especially until around 20% of the predicted event probability (Supplementary Figure 2).

**Figure 1.**
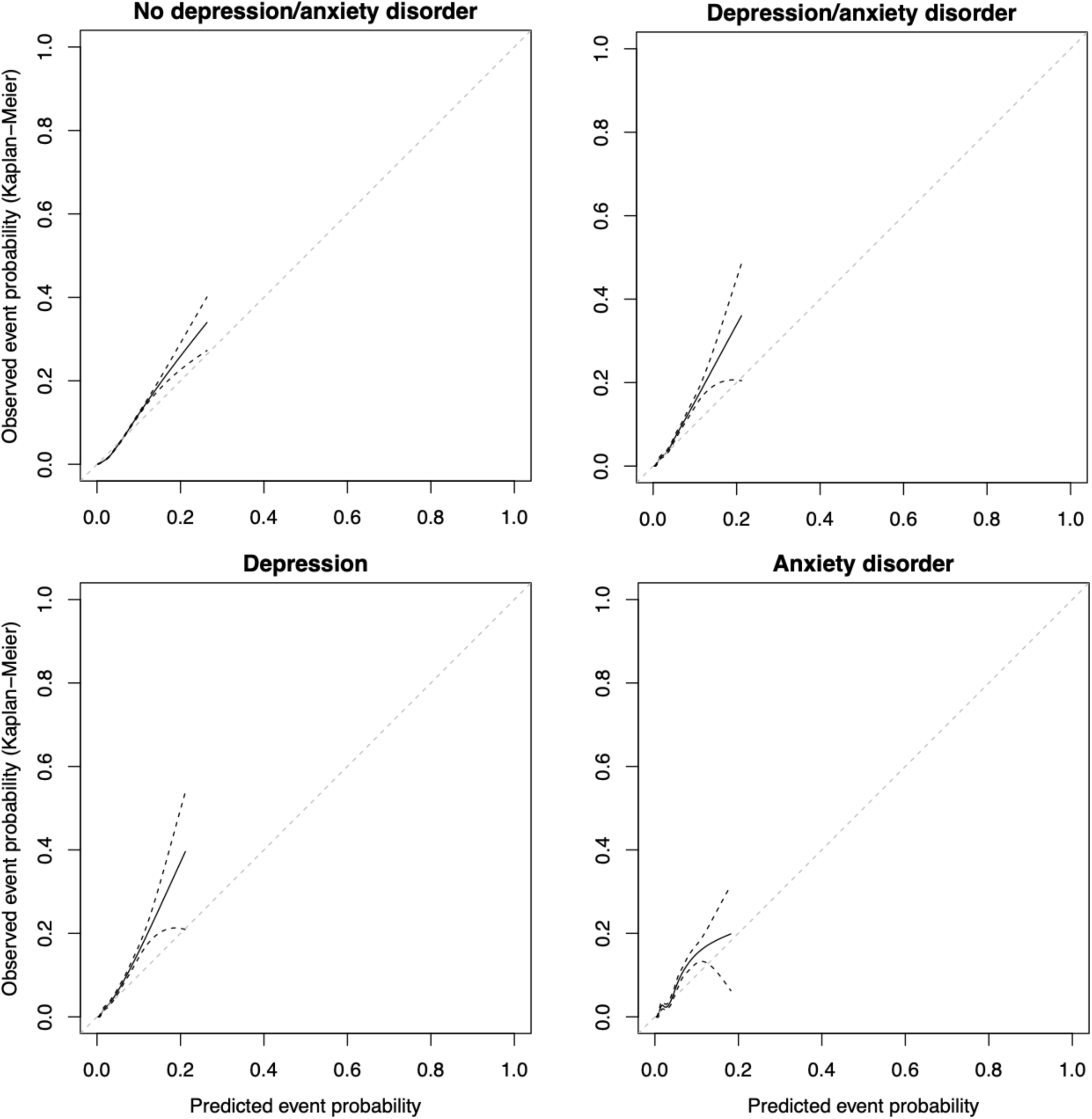
Calibration plots for 10-year cardiovascular disease risk among participants with and without depression and anxiety disorder

**Table 2.**
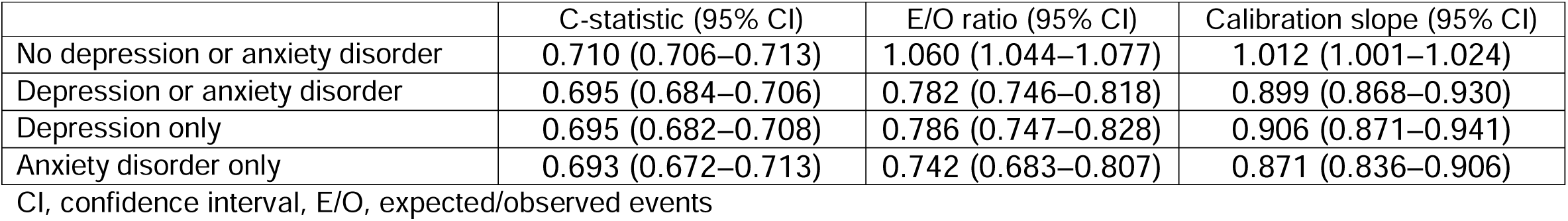
Performance of SCORE2 at predicting 10-year cardiovascular disease risk in participants with and without depression or anxiety der.

|  | C-statistic (95% CI) | E/O ratio (95% CI) | Calibration slope (95% CI) |
| --- | --- | --- | --- |
| No depression or anxiety disorder | 0.710 (0.706–0.713) | 1.060 (1.044–1.077) | 1.012 (1.001–1.024) |
| Depression or anxiety disorder | 0.695 (0.684–0.706) | 0.782 (0.746–0.818) | 0.899 (0.868–0.930) |
| Depression only | 0.695 (0.682–0.708) | 0.786 (0.747–0.828) | 0.906 (0.871–0.941) |
| Anxiety disorder only | 0.693 (0.672–0.713) | 0.742 (0.683–0.807) | 0.871 (0.836–0.906) |

CI, confidence interval, E/O, expected/observed events

After recalibration, 5,529 (38.0%) of those aged ≥50 years and 1,464 (23%) of those aged <50 years with these mental health conditions were reclassified upward to the intermediate or higher predicted risk category (Figure 2 and 3 and Supplementary Table 2 and 3). Dividing these by NNT by statins yielded up to 191 (11.7%) and 51 (27.4%) potentially preventable CVD cases among those aged ≥50 years and those aged <50 years, respectively (Supplementary Tables 4 and 5). Overall, sub-group analyses by depression and anxiety disorder produced consistent findings, whereby all measures indicated lower predictive performance and the recalibrated models yielded potentially preventable CVD cases.

**Figure 2.**
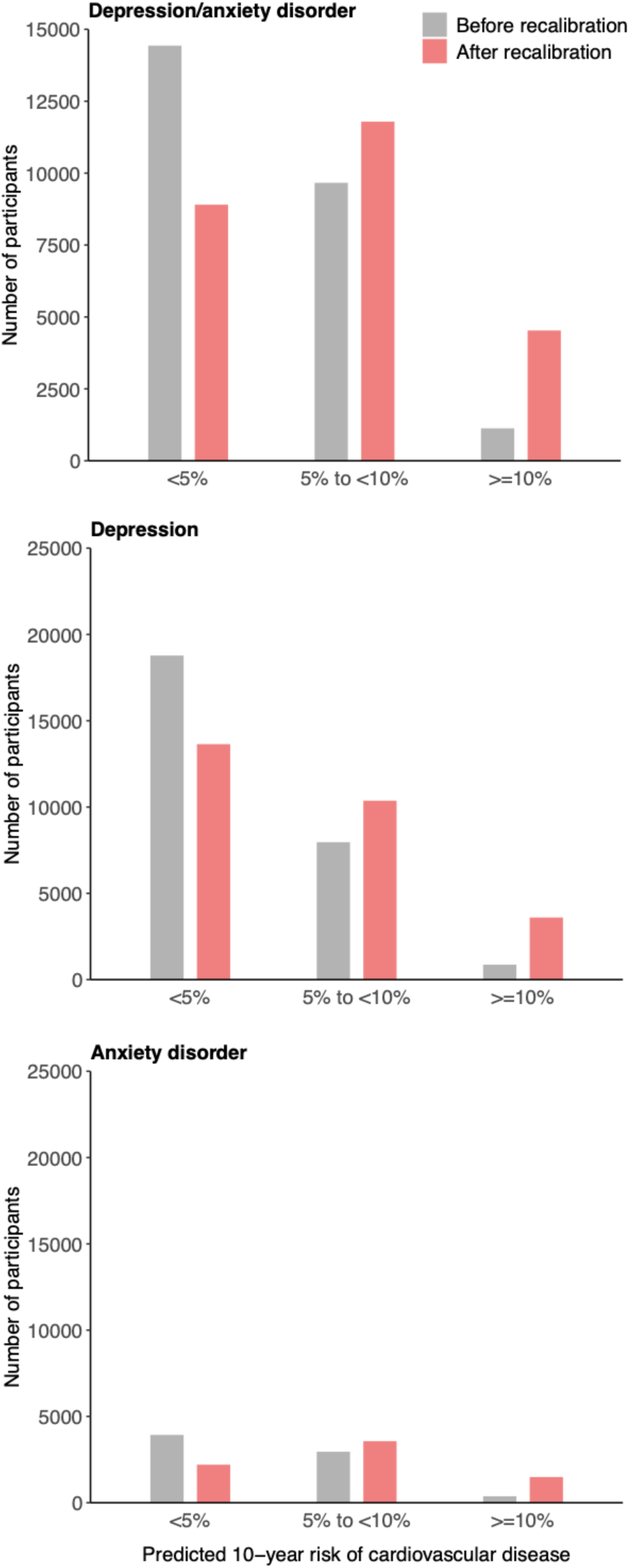
Numbers of participants assigned to low, intermediate, and high predicted 10-year cardiovascular disease risk categories before and after recalibration in the age group of ≥50 years

**Figure 3.**
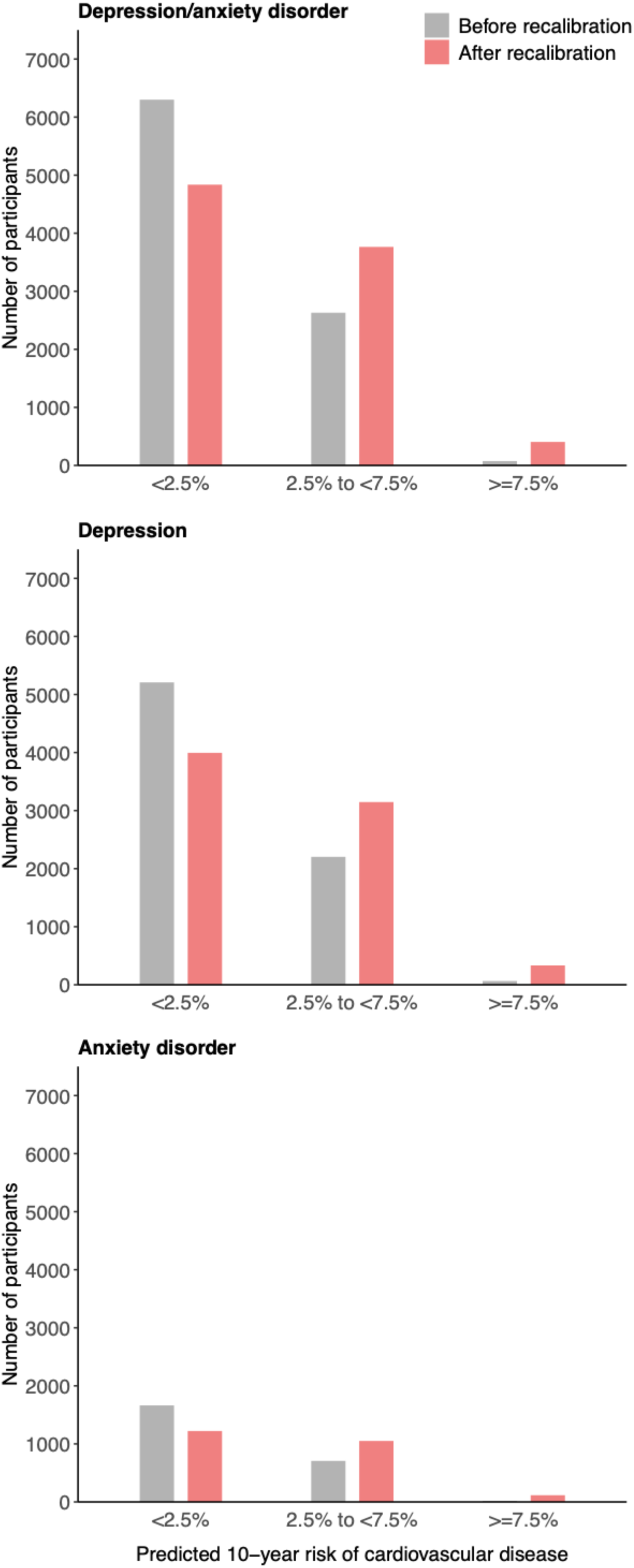
Numbers of participants assigned to low, intermediate, and high predicted 10-year risk categories of cardiovascular disease before and after recalibration in the age group of <50 years

The highest missing predictor variable was HDL-c (Supplementary Table 6), but the characteristics of participants included in the analysis and excluded due to missing values were similar in terms of the distributions of CVD risk factors and CVD events (Supplementary Table 7).

## Discussion

We demonstrated that SCORE2 underperformed at predicting 10-year CVD risk among those with depression or anxiety disorder in terms of both discrimination and calibration measures. In particular, calibration measures indicated a systematic underestimation of CVD risk. The analyses suggested that the recalibrated models could predict additional CVD cases that are potentially preventable by statin therapy irrespective of age group.

The observed risk underestimation among those with depression or anxiety disorder would be attributable to their distinct characteristics that SCORE2 could not capture. Although SCORE2 can capture the high prevalence of smoking in this population, it does not account for other CVD risk factors that are disproportionately prevalent. For example, accumulating studies have found higher inflammatory activity among those with these mental health conditions [9, 15]. Although we could not find any directly relevant studies, CVD risk scores could underperform at predicting the risk in specific populations, such as people with diabetes or schizophrenia [16, 17]. Indeed, specific risk scores were developed for these populations to address this issue [18, 19].

The use of SCORE2 is currently standardised (except for specific populations such as diabetes) [6], and it may lead to CVD events that could have been prevented with statin therapy among those with depression or anxiety disorder. However, this could be avoided by relatively simple recalibration without developing new models with new biomarkers requiring additional costs and invasive procedures. Notably, we confirmed that all results were consistent between those with depression and anxiety disorder. These two mental health conditions would have not only shared but also distinct characteristics [8, 20] that could lead to differential CVD risks. However, our results did not support the necessity of differentiating them when assessing CVD risk.

Our findings would apply to a wide spectrum of depression and anxiety disorder encompassing cases managed in the community and in hospital because we ascertained them through multiple sources, including self-report and primary care and hospital admission data. However, common mental health conditions can be overlooked when patients are treated mainly for other physical problems in primary care settings. In such cases, routine screening for these conditions could maximise the impact of our findings. In contrast to the USA, European countries do not have guidelines currently, nor does the UK National Screening Committee recommend routine depression screening [21]. Because the early detection and treatment of depression would improve health outcomes, the US Preventive Services Task Force has recommended routine screening for depression among adults in primary care settings [22].

There are several limitations to our study. Firstly, we used the UK Biobank cohort, which was included in the SCORE2 development cohorts. The SCORE2 performance in our study would be better than that in fully independent validation data, where participant characteristics would differ from those in the UK Biobank. Future studies should evaluate the SCORE2 performance using fully independent data on people with depression or anxiety disorder, especially from multiple European countries. Secondly, we identified prevalent depression and anxiety disorder using self-report of past medical history and historical clinical records and some historical cases may have already been in remission at the baseline. Therefore, our results could still underestimate the CVD risk among those who suffered mental health conditions at the time of CVD risk assessment. Thirdly, we excluded participants due to missing values in predictors. This resulted in an 18% sample size reduction and could introduce a bias in estimates if the included participants are no longer representative of the original sample. However, we confirmed the numbers of events and non-events well exceeded 100 with very little difference in characteristics between those included and excluded from the analyses. Lastly, we obtained the 10-year NNT values of relevant CVD outcomes from the West of Scotland Coronary Prevention Study, which recruited male participants with high LDL-c concentrations, implying high baseline CVD risk [23]. The NNT values in this trial would be lower, and consequently, the estimated preventable CVD cases would be higher than had we had access to data from other trials with lower CVD risk profiles, such as the AFCAPS/ TexCAPS and JUPITER trials [24, 25]. However, we could not use these trials due to the lack of 10-year follow-up data.

## Conclusions

SCORE2 could underestimate 10-year CVD risk among people with depression or anxiety disorder, potentially leading to CVD events that could have been prevented with statin therapy. Therefore, model recalibration is recommended for clinical application in this population.

## Data Availability

UK Biobank data can be requested by bona fide researchers for approved projects, including replication, through https://www.ukbiobank.ac.uk/.

## Acknowledgements

We are grateful to the UK Biobank participants. This research has been conducted using the UK Biobank Resource under application number 71392. We are also grateful to the Medical Research Council and the University of Edinburgh/University of Glasgow. This work was supported by the Medical Research Council (MR/N013166/1-LGH/MS/MED2525).

## Declaration of interests

The authors declare that there is no conflict of interest regarding the publication of this article.

## Author Contribution

SN: conceptualisation, formal analysis, writing – original draft CM: writing – review & editing

FH, JP: conceptualisation, writing – review & editing

## Funding

S.N. is supported by a Ph.D. studentship award from the Medical Research Council (MR/N013166/1-LGH/MS/MED2525).

## Notes

### Competing Interest Statement

The authors have declared no competing interest.

