## Supplemental Materials for "Predictive performance of SCORE2 among people with depression and anxiety disorder"

**Supplementary Materials for Predictive performance of SCORE2 among people with depression and anxiety disorder**

**Supplementary Figure 1.** Flowchart of the participant selection process

**Supplementary Figure 2.** Calibration plots for 10-year cardiovascular disease risk among participants with and without depression and anxiety disorder after recalibration

**Supplementary Table 1.** ICD10-codes used to define cardiovascular disease outcome

**Supplementary Table 2.** Numbers of participants assigned to low, intermediate, and high predicted 10-year risk categories of cardiovascular disease before and after recalibration in the age group ≥50 years

**Supplementary Table 3.** Numbers of participants assigned to low, intermediate, and high predicted 10-year risk categories of cardiovascular disease before and after recalibration in the age group <50 years

**Supplementary Table 4.** Additional 10-year cardiovascular disease cases potentially preventable by statin, derived from SCORE2 before and after recalibration in the age group of ≥50 years

**Supplementary Table 5.** Additional 10-year cardiovascular disease cases potentially preventable by statin, derived from SCORE2 before and after recalibration in the age group of <50 years

**Supplementary Table 6.** Summary of missing values in predictors

**Supplementary Table 7.** Comparisons of characteristics of participants who were included in the analysis and excluded from the analysis due to missing values in predictors

Supplementary Figure 1. Flowchart of the participant selection process

502,355 UK Biobank participants

486,720

15,646 excluded due to cardiovascular disease/diabetes history

401,453

86,267 excluded due to missing values in predictors

34,228	participants with depression/anxiety disorder

367,225 participants without depression/anxiety disorder

Supplementary Figure 2. Calibration plots for 10-year cardiovascular disease risk among participants with and without depression and anxiety disorder after recalibration

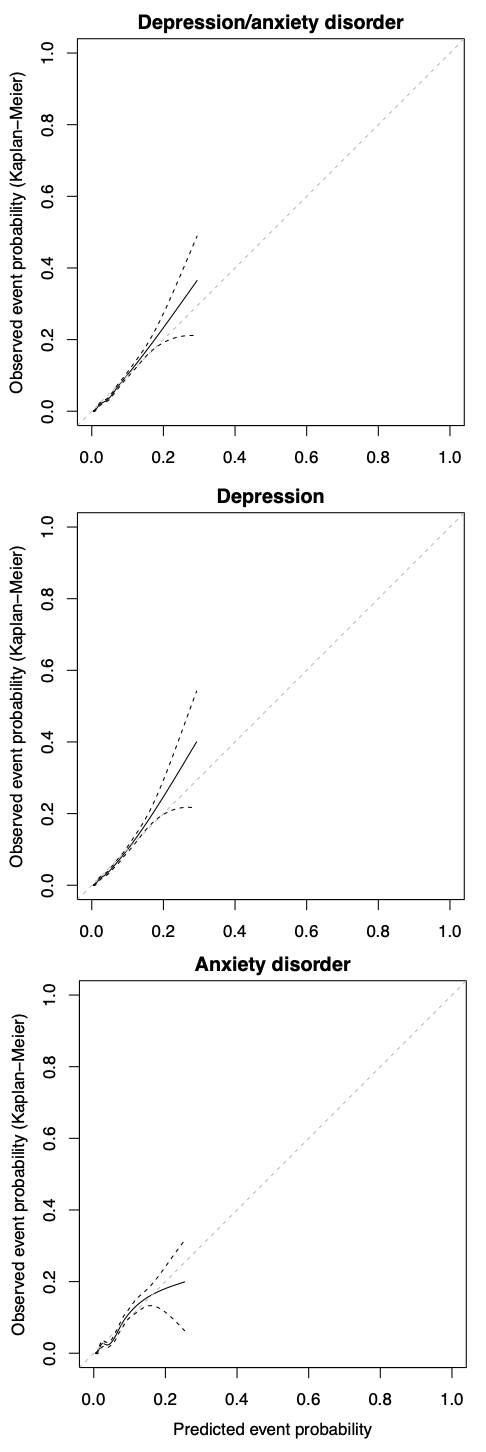

Supplementary Table 1. ICD10-codes used to define cardiovascular disease outcome

| Fatal cardiovascular disease | |
| --- | --- |
| ICD10-codes included in the outcome | ICD10-codes excluded from the outcome |
| Hypertensive disease: I10-16 |  |
| Ischemic heart disease: I20-25 |  |
| Arrhythmias, heart failure: I46-52 | I51.4 |
| Cerebrovascular disease: I60-69 | I60, I62, I67.1, I67.5, I68.2 |
| Atherosclerosis/AAA: I70-73 |  |
| Sudden death and death within 24h of symptom onset: R96.0-96.1 |  |
| Non-fatal cardiovascular disease | |
| ICD10-codes included in the outcome | ICD10-codes excluded from the outcome |
| Myocardial infarction: I21-I23 |  |
| Stroke: I60-69 | I60, I62, I67.1, I67.5, I68.2 |

Supplementary Table 2. Numbers of participants assigned to low, intermediate, and high predicted 10-year risk categories of cardiovascular disease before and after recalibration in the age group ≥50 years

|  | Number of participants | | |
| --- | --- | --- | --- |
|  | <5% | 5% to <10% | ≥10% |
| **Depression/anxiety disorder** |  |  |  |
| Before recalibration | 14,432 | 9,664 | 1,127 |
| After recalibration | 8,903 | 11,790 | 4,530 |
| **Depression** |  |  |  |
| Before recalibration | 11,710 | 7,566 | 858 |
| After recalibration | 7,234 | 9,395 | 3,505 |
| **Anxiety disorder** |  |  |  |
| Before recalibration | 3,928 | 2,955 | 375 |
| After recalibration | 2,204 | 3,566 | 1,488 |

Supplementary Table 3. Numbers of participants assigned to low, intermediate, and high predicted 10-year risk categories of cardiovascular disease before and after recalibration in the age group <50 years

|  | Number of participants | | |
| --- | --- | --- | --- |
|  | <2.5% | 2.5% to <7.5% | ≥7.5% |
| **Depression/anxiety disorder** |  |  |  |
| Before recalibration | 6,300 | 2,630 | 75 |
| After recalibration | 4,836 | 3,764 | 405 |
| **Depression** |  |  |  |
| Before recalibration | 5,208 | 2,203 | 64 |
| After recalibration | 3,995 | 3,147 | 333 |
| **Anxiety disorder** |  |  |  |
| Before recalibration | 1,663 | 707 | 19 |
| After recalibration | 1222 | 1051 | 116 |

Supplementary Table 4. Additional 10-year cardiovascular disease cases potentially preventable by statin, derived from SCORE2 before and after recalibration in the age group of ≥50 years

|  | Additional preventable cases (%) | | |
| --- | --- | --- | --- |
|  | cardiovascular disease death  10-year NNT: 125 | Incident coronary heart disease 10-year NNT: 29 | Incident stroke  10-year NNT: 200 |
| Depression or anxiety disorder | 44.2 (2.7) | 191 (11.7) | 27.6 (1.7) |
| Depression only | 35.8 (2.8) | 154 (12.0) | 22.4 (1.7) |
| Anxiety disorder only | 13.8 (2.7) | 59.4 (11.8) | 8.62 (1.7) |

NNT, Number Needed to Treat; NA, not available

The additional preventable cases were calculated as the difference in the number of participants in risk groups (≥5%) between before and after recalibration models divided by 10-year NNT. 10-year NNT was obtained from West of Scotland Coronary Prevention Study. The proportion (%) was calculated by dividing the additional preventable cases by the observed cardiovascular disease cases.

Supplementary Table 5. Additional 10-year cardiovascular disease cases potentially preventable by statin, derived from SCORE2 before and after recalibration in the age group of <50 years

|  | Additional preventable cases (%) | | |
| --- | --- | --- | --- |
|  | cardiovascular disease death  10-year NNT: 125 | Incident coronary heart disease 10-year NNT: 29 | Incident stroke  10-year NNT: 200 |
| Depression or anxiety disorder | 11.7 (6.4) | 50.5 (27.4) | 7.3 (4.0) |
| Depression only | 9.7 (6.6) | 41.8 (28.4) | 6.1 (4.1) |
| Anxiety disorder only | 3.5 (6.3) | 15.2 (27.6) | 2.2 (4.0) |

NNT, Number Needed to Treat; NA, not available

The additional preventable cases were calculated as the difference in the number of participants in risk groups (≥2.5%) between before and after recalibration models divided by 10-year NNT. 10-year NNT was obtained from West of Scotland Coronary Prevention Study. The proportion (%) was calculated by dividing the additional preventable cases by the observed cardiovascular disease cases.

Supplementary Table 6. Summary of missing values in predictors

| Predictors | N (%) |
| --- | --- |
| High density lipoprotein cholesterol | 70,274 (14.4) |
| Total cholesterol | 31,794 (6.5) |
| Systolic blood pressure | 16,242 (3.3) |
| Smoking | 2,767 (0.6) |
| Age | 15 (0.003) |
| Sex | 15 (0.003) |

N, number

Supplementary Table 7. Comparisons of characteristics of participants who were included in the analysis and excluded from the analysis due to missing values in predictors

|  | Participants included in the analysis | | | Participants excluded from the analysis due to missing values in predictors | | |
| --- | --- | --- | --- | --- | --- | --- |
|  | Total | Non-CVD | CVD | Total | Non-CVD | CVD |
|  | N = 401,453 | N = 371,022 | N = 30,431 | N = 85,267 | N = 78,306 | N = 6,961 |
| Age (years), mean (SD) | 56.4 (8.10) | 56.0 (8.08) | 61.2 (6.67) | 56.3 (8.13) | 55.9 (8.10) | 61.1 (6.77) |
| Smoking, N (%) | 42,122 (10.49) | 37,014 (9.98) | 5,108 (16.79) | 8,810 (10.68) | 7,648 (10.08) | 1,162 (17.50) |
| Systolic blood pressure (mmHg), mean (SD) | 137.9 (18.60) | 137.3 (18.41) | 144.8 (19.47) | 137.5 (18.87) | 136.8 (18.62) | 144.7 (20.20) |
| Total cholesterol (mmol/l), mean (SD) | 5.7 (1.12) | 5.7 (1.11) | 5.6 (1.24) | 5.7 (1.14) | 5.7 (1.12) | 5.6 (1.28) |
| HDL-c (mmol/l), mean (SD) | 1.5 (0.38) | 1.5 (0.38) | 1.4 (0.38) | 1.4 (0.38) | 1.4 (0.38) | 1.3 (0.38) |

CVD, cardiovascular disease; N, number; SD, standard deviation; HDL-c, high density lipoprotein cholesterol
